# Increased HIV incidence during Event-Driven PrEP compared to Daily PrEP in the Netherlands

**DOI:** 10.64898/2026.08.12.26360235

**Authors:** I.J.M. Willemstein, M. Prins, J.C.M. Heijne, U. Davidovich, M.F. Schim van der Loeff, L. Chanamé Pinedo, E.U. Akwiwu, B.H.B. van Benthem, E. Hoornenborg, V.W. Jongen

**Affiliations:** Department of Infectious Diseases, Public Health Service Amsterdam, Amsterdam, the Netherlands; Amsterdam UMC, Univ of Amsterdam, Internal Medicine, Amsterdam institute of Immunology and Infectious Diseases (AI&I), and Amsterdam Public Health research institute, Meibergdreef 9, Amsterdam, the Netherlands; University of Amsterdam, Department of Social Psychology, Amsterdam, The Netherlands; Centre for Infectious Disease control, National Institute for Public Health and the Environment, Bilthoven, The Netherlands; Amsterdam UMC, Vrije Universiteit Amsterdam, Department of Epidemiology and Data Science, Amsterdam Public Health, Amsterdam, The Netherlands; Stichting hiv monitoring, Amsterdam, The Netherlands

## Abstract

**Background:** Clinical trials demonstrated high efficacy of daily and event-driven oral pre-exposure prophylaxis (PrEP) in HIV prevention. Event-driven PrEP involves taking two tablets before and two times one tablet after sexual contact (2-1-1/on-demand). While both are implemented in Dutch clinical practice, evaluating real-world effectiveness requires large-scale data from routine clinical care. This study compared HIV incidence between daily and event-driven PrEP in the Netherlands.

**Methods:** We used surveillance data from the Dutch national PrEP program (August 1, 2019-December 31, 2025). Individuals ≥16 years with ≥1 follow-up consultation after PrEP initiation were included; PrEP regimen since last visit was recorded at each visit. Person-time was modeled as time-varying based on the regimen reported at each consultation. HIV incidence rates were calculated per 100 person-years and Cox proportional hazards models estimated hazard ratios between regimens for HIV acquisition, adjusted for sociodemographics, sexual behavior, and history of sexually transmissible infections.

**Findings:** 16,469 individuals (15,843 men who have sex with men, 579 transgender and gender diverse persons, 45 women and two men who have sex with women) initiated PrEP and had ≥1 follow-up visit (median follow-up 2·0 years (IQR=0·8-4·0)). Median age was 33 years (IQR=27-44). 49 PrEP users were diagnosed with HIV over 41,092 person-years (IR=0·12/100 py;95%CI=0·09-0·16), of whom 42 event-driven users (IR=0·20/100 py;95%CI=0·15-0·27) and seven daily PrEP users (IR=0·04/100 py;95%CI=0·02-0·07). In multivariable Cox regression, event-driven PrEP use was associated with a higher hazard of HIV acquisition (aHR=7·0;95%CI=3·0-16·4).

**Interpretation:** Despite overall low HIV incidence, the incidence rate in the Dutch national PrEP program was seven-fold higher during event-driven PrEP use compared to daily, which may be due to lower adherence. These findings denotes that, in real-world settings, improved person-centered counseling is needed for individuals interested in, or using event-driven PrEP. Research should identify domains and preferred methods of support.

**Funding:** None for this study.

**Research in context:** *Evidence before this study:* We searched PubMed on 07 July 2026 without language or date restrictions, using the (Mesh)terms (“HIV”[Mesh] OR hiv OR hiv-1 OR hiv-2 OR hiv infections OR acquired immunodeficiency syndrome OR (human immunodeficiency virus* OR human immunodeficiency virus* OR human immuno-deficiency virus* OR human immune-deficiency virus*)) AND (“Incidence”[Mesh] OR incidence) AND (“Pre-Exposure Prophylaxis”[Mesh] OR “Tenofovir”[Mesh] OR “Emtricitabine”[Mesh] OR (“PrEP” OR “Pre-Exposure Prophylaxis” OR “Preexposure Prophylaxis” OR “HIV Pre-Exposure Prophylaxis” OR “HIV PrEP” OR truvada OR emtricitabine OR tenofovir)) AND (event-driven OR intermittent OR on-demand OR 2-1-1 OR “on demand”). This search yielded 213 publications. Clinical trials and cohort studies demonstrated low HIV incidence across both daily and event-driven oral tenofovir disoproxil / emtricitabine (TDF-FTC) PrEP regimens. Most comparative real-world data originated from implementation and demonstration projects in the early oral PrEP era (2014-2020), with follow-up periods ranging from six months to four years. To date, only the ANRS PREVENIR study (a prospective cohort study across 22 sites in the Paris region) provided more recent data with longer follow-up time (i.e., a seven-year follow-up period up to May 2024). Despite high efficacy and effectiveness in study settings, there is evidence that in real-life some event-driven PrEP users incorrectly use PrEP or have more difficulties adhering to the PrEP regimen in comparison to daily PrEP users, in particular young men who have sex with men. A few studies have found a higher HIV incidence among event-driven oral PrEP users than among daily oral PrEP users, related to lower adherence, but these findings were derived from small-scale cohort studies with a low number of participants and restricted to a limited number of clinical sites rather than a national cohort.

*Added value of this study:* Using real-world national surveillance data comprising data over six years since nationwide implementation of the Dutch national PrEP programme, we evaluated HIV incidence by PrEP regimen. While the overall HIV incidence was low, we found a seven times higher HIV incidence among individuals who reported having used event-driven oral PrEP compared to those reporting daily oral PrEP use, after adjusting for socio-demographics, sexual health and behavior. To the best of our knowledge, this is the first study reporting a significant difference in HIV incidence between the two oral regimens using national surveillance data over an extended follow-up period.

*Implications of all the available evidence:* While event-driven oral PrEP has been demonstrated to be as effective as daily use in study settings, most likely related to good adherence, individual and contextual challenges might have led to insufficient adherence resulting in lower effectiveness in a real-world setting. This underscores the need to better understand the real-world challenges relating to event-driven PrEP use and improved person-centered counseling of event-driven PrEP.

## INTRODUCTION

Clinical trials demonstrated the efficacy of oral pre-exposure prophylaxis (PrEP) in preventing HIV acquisition when taken as a daily or event-driven (also known as on-demand or intermittent) regimen (i.e., two tablets between 24 and two hours before sex, followed by two times one tablet after 24 and 48 hours).^1,2^ Both regimens are recommended by the World Health Organization (WHO) to offer individuals flexibility, choice, convenience, and prevention tailored to their needs.^3^

In the Netherlands, both regimens are included in national PrEP guidelines, and available through general practitioners and the Dutch national PrEP program since 2019. The national PrEP program is implemented at all Centers for Sexual Health throughout the Netherlands, providing PrEP care to individuals who can benefit from PrEP the most, including men who have sex with men (MSM), transgender and gender diverse persons (TGDP), and sex workers.^4^ Since the start of the national program, an increasing proportion of PrEP users has used event-driven PrEP, with event-driven PrEP use being reported in half of all PrEP consultations at Centers for Sexual Health in 2025.^5^ There is, however, some evidence that the effectiveness of event-driven PrEP may be reduced compared to daily PrEP due to incorrect usage or adherence issues.^6–8^

Given the potential implications of possible reduced effectiveness of event-driven PrEP, and programmatic implications regarding enhanced adherence counselling, there is a need to better examine the real-world effectiveness of event-driven PrEP. However, longitudinal real-world data on HIV incidence stratified by PrEP regimen are scarce. Therefore, we aimed to compare HIV incidence during daily and event-driven PrEP use over six years after implementation of the Dutch national PrEP program.

## METHODS

### Study design and population

We conducted a retrospective cohort study using surveillance data from the Dutch national PrEP program implemented at all Centers for Sexual Health in the Netherlands (datasource: SOAP; Seksueel Overdraagbare Aandoeningen Peilstation) from August 1, 2019, to December 31, 2025. Individuals aged ≥16 years old who were PrEP users and who had at least one follow-up consultation after PrEP initiation were included in this study.

### Setting

Oral PrEP, in the form of tenofovir disoproxil 245mg / emtricitabine 200mg tablets, has been available in the Netherlands since August 2019 via a national program encompassing 23 Centers for Sexual Health at Public Health Services. In addition PrEP is provided through general practitioners located throughout the Netherlands. For this analysis, we used data from the Centers for Sexual Health. Initially, PrEP was available through a nationally coordinated pilot program (2019-2024), capped at 8,500 people at increased chance for HIV acquisition. Eligibility criteria for PrEP were having had at least one of the following in the past six months: insertive and/or receptive anal sex without a condom, anal STI or infectious syphilis, post-exposure prophylaxis (PEP) use; problematic chemsex use was added as a criterion in 2022, based on a clinician’s assessment.^4^ During this national pilot, the standard visit frequency was three-monthly and PrEP tablets could be obtained in the clinic at reduced costs (€7.50 per 30 tablets). In August 2024, the pilot program ended and PrEP care was structurally integrated into routine care at Centers for Sexual Health. Since then, the standard visit frequency has been extended to four-to six-monthly, based on the clinician’s assessment. PrEP tablets were obtained through public pharmacies with costs ranging from €15 to €65 per 30 tablets, depending on the pharmacy.

Centers for Sexual Health provide free-of-charge consultations including testing for sexually transmitted infections (STI; i.e. chlamydia, gonorrhea, syphilis), HIV, hepatitis B, hepatitis C, kidney function assessment and vaccination for hepatitis B and mpox. At the initial PrEP visit, all individuals undergo comprehensive screening, including an HIV test. Immediate PrEP initiation is facilitated. At each follow-up visit, PrEP users are tested for bacterial STI, HIV and are offered face-to-face counseling. Individuals could attend the clinic for STI testing in-between PrEP visits.

### Outcomes and variables

The main outcome in this study was HIV incidence, stratified by PrEP regimen. Secondary outcomes were discontinuation incidence and loss-to-follow-up (LTFU) incidence, estimated separately per PrEP regimen. PrEP discontinuation was defined as people who formally notified the Center of Sexual Health about discontinuing PrEP and LTFU as no visits within nine months since last PrEP consultation. PrEP use and regimen in the past year, specified for the past three months and earlier periods, was reported at each PrEP visit. For the main analysis, PrEP use reported in the past three months was used, while consultations where PrEP use was reported between 12 and four months ago were excluded. A sensitivity analysis additionally included this period. For PrEP initiation visits, we assumed the individual had initiated PrEP using the regimen reported at the following visit. In a sensitivity analysis, we assumed all individuals had initiated PrEP using the daily regimen. This assumption was based on clinical practice where individuals are typically advised to start with daily PrEP for the initial month.

Other variables included were age (continuous), sexual preference (categorized as MSM/other), education level (tertiary, upper secondary/vocational, lower secondary/preparatory), migration background (no, second or first generation migration background), STI diagnosis history in the past year (i.e., chlamydia, gonorrhea, syphilis), and the following behavioral variables in the past six months: number of sexual partners (0-4, 5-9, 10+), condomless anal sex (CAS) (yes/no), sex work (yes/no), group sex (yes/no), chemsex (yes/no, the use of methamphetamine, mephedrone, GHB/GBL, ketamine, cocaine, amphetamine/speed, 3-MMC, or any new psychoactive substances before or during sex). For socio-demographic variables (i.e., age, sexual preference, education level, migration background), missing values were supplemented using non-missing data from a subsequent consultation where available. This was not applied to behavioral variables due to their time-varying nature.

### Statistical analysis

We described characteristics at first PrEP consultation, overall and stratified by initial PrEP regimen (i.e., daily or event-driven PrEP), as reported at the first follow-up visit. Differences between groups were assessed using the Wilcoxon rank sum test for continuous variables and Pearson’s chi-square test or Fisher’s exact test for categorical variables.

First, we calculated discontinuation rates after periods of daily and event-driven PrEP. Specifically, incidence rates of formal discontinuation were estimated per 100 person-years (py) using Poisson regression with the logarithm of person-years as an offset. For both outcomes, robust standard errors were used to account for within-individual correlation, as individuals could contribute multiple observations. Rates were computed separately for event-driven and daily PrEP regimens (i.e., this analysis allowed people to contribute to both regimens). Second, we calculated HIV incidence rates using the same approach as for discontinuation rates. Time at risk started at first PrEP consultation and ended at HIV diagnosis, PrEP discontinuation, LTFU (i.e., date of last visit prior to ≥9-month gap between consultations) or censoring date (i.e., December 31, 2025), whichever occurred first. For individuals diagnosed with HIV, we used the midpoint between the last negative and first positive test as the estimated date of seroconversion. For individuals who were LTFU, time-at-risk stopped at their last visit. Individuals could re-enter in the analyses if they returned for PrEP consultations; person-time re-started at the moment of re-entry. HIV diagnoses made at re-entry were considered prevalent and were thus excluded from the incidence analyses.

We estimated the descriptive cumulative HIV incidence (1-Kaplan-Meier survival) with PrEP regimen treated as time-varying exposure. Individuals who switched PrEP regimen contributed follow-up under their regimen until the time of switching and subsequently entered the risk set for the new regimen through delayed entry (left truncation). In case of missing data on PrEP regimen during follow-up, we imputed missing data within an individual using information from their previous consultation (last observation carried forward). In sensitivity analyses, we evaluated alternative approaches for handling missing data on PrEP regimen, including complete case analysis and last observation carried backwards.

We used a Cox proportional hazards model to evaluate the association between PrEP regimen and HIV acquisition, estimating hazard ratios (HRs) and their corresponding 95% confidence intervals (CIs). Based on a-priori selection, we adjusted the Cox proportional hazards model for all previously specified sociodemographic factors, STI history, and behavioral characteristics. Migration background and education level were included as time-fixed covariates; sexual preference, sexual health and behavior as time-varying covariates. Missing data were analyzed as a separate category. Exceptions were made when an ‘unknown’ category would be non-informative as it contained no HIV diagnoses. Specifically, for migration background, missings were kept as missing and excluded from the model. For STI diagnosis history, sexual preference and gender, missing values were collapsed into the reference categories (i.e., STI history vs. no STI history/unknown and ‘Woman/Men who have Sex with Women(MSW)/TGDP/Unknown vs. MSM, respectively). Through these approaches we aimed to minimize exclusion of observations. Based on the a priori hypothesis that behavior and PrEP regimen interact in their association with HIV acquisition, interaction terms were first assessed in unadjusted models before fitting the multivariable models. Due to the limited number of events, full multivariable models incorporating interaction terms could not be fitted to avoid overfitting. To correct for within-person correlation, robust standard errors were used. We assessed the proportional hazards assumption using Schoenfeld residuals and visual inspection of log-minus-log survival plots. Finally, we assessed multicollinearity among covariates using the Variance Inflation Factor (VIF). In cases of high colinear variables (VIF>10), the variable with the strongest confounding effect was retained in the analysis.

All analyses were conducted using RStudio, version 4.5.1 (packages survival, survminer).

### Ethics approval

Individuals visiting Centers for Sexual Health can opt-out of sharing their data. SOAP data may be used for research purposes after approval from the registration committee. The database is provided in pseudonymized form. Under Dutch national law, for retrospective studies using routinely collected surveillance data additional ethical review and informed consent are not required.

## RESULTS

Between August 1, 2019, and December 31, 2025, 18,047 people had an initial PrEP consultation and reported using PrEP during the study period. After excluding 1,578 individuals who lacked follow-up consultations, or were diagnosed with HIV at their initial PrEP consultation (n=5), 16,469 (91·3%) remained for further analyses. In total, these 16,469 individuals accounted for 144,946 consultations: 123,520 (85·2%) for PrEP and 21,426 (14·8%) for in-between STI testing.

Of these 16,469 PrEP users, 8,330 (50·6%) started daily PrEP use at baseline and 8,139 (49·4%) event-driven PrEP (Table 1). The proportion event-driven PrEP increased over time from 37.0% (344/930) in 2019 to 61.2% (1,733/2,832) in 2025. 15,843 (96·2%) were MSM, 579 (3·5%) TGDP, 45 (0·3%) women and two (0·0%) men who have sex with women. Median age was 33 years (IQR=27-44), 10,405 (63·2%) had completed a tertiary education and 7,515 (45·6%) had a first- or second-generation migration background. In the preceding six months, median number of sex partners was six (IQR=3-12), condomless anal sex was reported by 14,510 individuals (88·1%), group sex by 5,782 (35·1%), chemsex by 1,295 (7·9%), and sex work by 775 (4·7%). 7,258 (44·1%) individuals were diagnosed with at least one STI in the preceding year.

**Table 1.** Baseline characteristics of 16,469 PrEP users in the national PrEP program, overall and by initiated PrEP regimen, the Netherlands, August 2019-December 2025.

| Baseline characteristics | Total<br>n = 16469<br>n (%) | Daily PrEP<br>n = 8330<br>n (%) | Event-driven PrEP<br>n = 8139<br>n (%) | P-value |
| --- | --- | --- | --- | --- |
| **Age (years), median [IQR]** | 33 (27–44) | 34 (28–44) | 33 (27–43) | **<0.001** |
| **Sexual preference and gender(a)** |  |  |  | **<0.001** |
| Women | 45 (0.3%) | 39 (0.5%) | 6 (0.1%) |  |
| MSW | 2 (0.0%) | 2 (0.0%) | 0 (0.0%) |  |
| MSM | 15,843 (96.2%) | 7,896 (94.8%) | 7,947 (97.6%) |  |
| TGDP | 579 (3.5%) | 395 (4.7%) | 184 (2.3%) |  |
| **Education level** |  |  |  | **<0.001** |
| Tertiary | 10,405 (63.2%) | 5,087 (61.1%) | 5,318 (65.3%) |  |
| Upper secondary/vocational | 3,391 (20.6%) | 1,738 (20.9%) | 1,653 (20.3%) |  |
| Lower secondary/preparatory | 1,336 (8.1%) | 730 (8.8%) | 606 (7.4%) |  |
| Unknown | 1,337 (8.1%) | 775 (9.3%) | 562 (6.9%) |  |
| **Migration background** |  |  |  | **<0.001** |
| No migration background | 8,944 (54.3%) | 4,294 (51.5%) | 4,650 (57.1%) |  |
| First generation migration background | 5,879 (35.7%) | 3,250 (39.0%) | 2,629 (32.3%) |  |
| Second generation migration background | 1,636 (9.9%) | 781 (9.4%) | 855 (10.5%) |  |
| Unknown migration background | 10 (0.1%) | 5 (0.1%) | 5 (0.1%) |  |
| **Number of partners(b), median [IQR]** | 6 (3–12) | 7 (4–15) | 5 (3–10) | **<0.001** |
| **Condomless anal sex(b)** |  |  |  | **0.014** |
| No | 1,733 (10.5%) | 825 (9.9%) | 908 (11.2%) |  |
| Yes | 14,510 (88.1%) | 7,366 (88.4%) | 7,144 (87.8%) |  |
| Unknown | 226 (1.4%) | 139 (1.7%) | 87 (1.1%) |  |
| **Group sex(b)** |  |  |  | **<0.001** |
| No | 8,297 (50.4%) | 3,887 (46.7%) | 4,410 (54.2%) |  |
| Yes | 5,782 (35.1%) | 2,966 (35.6%) | 2,816 (34.6%) |  |
| Unknown | 2,390 (14.5%) | 1,477 (17.7%) | 913 (11.2%) |  |
| **Chemsex(b,c)** |  |  |  | **<0.001** |
| No | 14,977 (90.9%) | 7,681 (92.2%) | 7,296 (89.6%) |  |
| Yes | 1,295 (7.9%) | 545 (6.5%) | 750 (9.2%) |  |
| Unknown | 197 (1.2%) | 104 (1.2%) | 93 (1.1%) |  |
| **Sex work(b)** |  |  |  | **<0.001** |
| No | 15,209 (92.3%) | 7,517 (90.2%) | 7,692 (94.5%) |  |
| Yes | 775 (4.7%) | 542 (6.5%) | 233 (2.9%) |  |
| Unknown | 485 (2.9%) | 271 (3.3%) | 214 (2.6%) |  |
| **STI diagnosis in past year** |  |  |  | **<0.001** |
| No | 7,596 (46.1%) | 3,593 (43.1%) | 4,003 (49.2%) |  |
| Yes | 7,258 (44.1%) | 3,910 (46.9%) | 3,348 (41.1%) |  |
| Unknown | 1,615 (9.8%) | 827 (9.9%) | 788 (9.7%) |  |
| **Year (first PrEP initiation visit)** |  |  |  | **<0.001** |
| 2019 | 930 (5.6%) | 586 (7.0%) | 344 (4.2%) |  |
| 2020 | 3,615 (22.0%) | 2,137 (25.7%) | 1,478 (18.2%) |  |
| 2021 | 3,103 (18.8%) | 1,602 (19.2%) | 1,501 (18.4%) |  |
| 2022 | 2,366 (14.4%) | 1,182 (14.2%) | 1,184 (14.5%) |  |
| 2023 | 1,898 (11.5%) | 916 (11.0%) | 982 (12.1%) |  |
| 2024 | 1,725 (10.5%) | 808 (9.7%) | 917 (11.3%) |  |
| 2025 | 2,832 (17.2%) | 1,099 (13.2%) | 1,733 (21.3%) |  |
| **Follow-up (years), median [IQR](d)** | **2.0 (0.8–4.0)** | **2.7 (1.1–4.5)** | **2.3 (1.0–4.1)** | **0.232** |

(b) In previous six months.
(c) Defined as the use of methamphetamine, mephedrone, GHB/GBL, ketamine, cocaine, amphetamine/speed, 3-MMC, or any new psychoactive substances before or during sex.
(d) Subgroup medians are higher because individuals could contribute to both PrEP regimes during their total follow-up.
IQR, Inter Quartile Range; MSW, Men who have Sex with Women; MSM, Men who have Sex with Men; TGDP, Transgender and Gender Diverse Persons; STI; Sexually Transmitted Infection.

Median follow-up time was 2·0 years (IQR=0·8-4·0) and median number of visits was seven (IQR=2-14). Based on the regimen reported at the last consultation, LTFU occurred after 496 daily and 1,213 event-driven PrEP regimen periods (IR=2·49/100py; 95%CI=2·28-2·73 and IR=5·72/100py; 95%CI=5·40-6·06, respectively). PrEP discontinuation occurred after 246 daily and 401 event-driven PrEP regimen periods (IR=1·24/100py; 95% CI=1·09·1-1·41 and IR=1·89/100py; 95% CI=1·71-2·09) (Table 2). After LTFU, 387/496 (78·0%) people who most recently used daily and 901/1,213 (74·3%) who used event-driven PrEP re-entered the PrEP program. After formal PrEP discontinuation, 65/246 (26·4%) people who most recently used daily PrEP and 96/401 (23·9%) who used event-driven PrEP re-entered. Eight people were diagnosed with HIV at re-entry; five had reported event-driven PrEP use at the last consultation before discontinuation or LTFU. These diagnoses were not included in HIV incidence analyses.

**Table 2.** Rates of official PrEP discontinuation, loss-to-follow-up and re-entry among PrEP users in the Dutch national PrEP program between August 1, 2019, and December 31, 2025.

|  | PrEP regimen | Individuals included | Person-years at risk | Individuals with event | Events | Incidence rate per 100 py (95% CI) | Incidence rate ratio (95% CI) | Individuals re-entered (%) |
| --- | --- | --- | --- | --- | --- | --- | --- | --- |
| Loss-to-follow-up | Daily PrEP | 10,687 | 19,881 | 489 | 496 | 2.49 (2.28-2.73) | REF | 387 (78.0%) |
| Loss-to-follow-up | Event-driven PrEP | 12,304 | 21,211 | 1138 | 1213 | 5.72 (5.40-6.06) | 2.29 (2.06-2.55) | 901 (74.3%) |
| PrEP discontinuation | Daily PrEP | 10,687 | 19,881 | 241 | 246 | 1.24 (1.09-1.41) | REF | 65 (26.4%) |
| PrEP discontinuation | Event-driven PrEP | 12,304 | 21,211 | 392 | 401 | 1.89 (1.71-2.09) | 1.53 (1.30-1.80) | 96 (23.9%) |

Note: PrEP regimen refers to the regimen at last consultation at time of PrEP discontinuation or loss-to-follow-up event. py, person-years; CI, confidence interval

A total of 49 individuals acquired HIV over 41,092 person-years of follow-up (IR=0·12/100py; 95%CI=0·09-0·16). When stratified by PrEP regimen, 42 people over 21,211 person-years who most recently used event-driven PrEP were diagnosed with HIV (IR=0·20/100py; 95%CI=0·15-0·27) and seven people over 19,881 person-years who most recently used daily PrEP (IR=0·04/100py; 95%CI=0·02-0·17). Of the 49 individuals who acquired HIV, 43 (87·8%) were MSM and six (12·2%) TGDP. Median age at HIV diagnosis was 33 years (IQR=29-48) and they reported a median number of five sex partners in the six months before HIV diagnosis (IQR=3-10).

Figure 1 shows the difference in cumulative HIV incidence during daily and event-driven PrEP use. Consistent with these survival curves, the univariable Cox proportional hazards model showed that event-driven PrEP was associated with a significantly higher hazard of HIV acquisition than daily PrEP (HR=5·6, 95%CI=2·5-12·5; p<0·001) (Table 3). This association remained significant in the multivariable model (aHR=7·0, 95%CI=3·0-16·4; p<0·001), as well as the sensitivity analysis also including PrEP use between 12 and 4 months ago (aHR=4·3, 95%CI=2·1-8·7; p<0·001) (Supplementary Material S2-S3). Unadjusted models yielded no statistically significant interactions, with some terms being not estimable due to zero numbers of events (Supplementary Table S4). Consequently, no interaction terms were included in the final adjusted model. Sensitivity analyses using alternative approaches to handle missing data yielded results similar to the main analysis (Supplementary Material S5). The same holds for sensitivity analyses assuming that all individuals initiated daily PrEP (Supplementary Material S6-S7).

**Figure 1.**
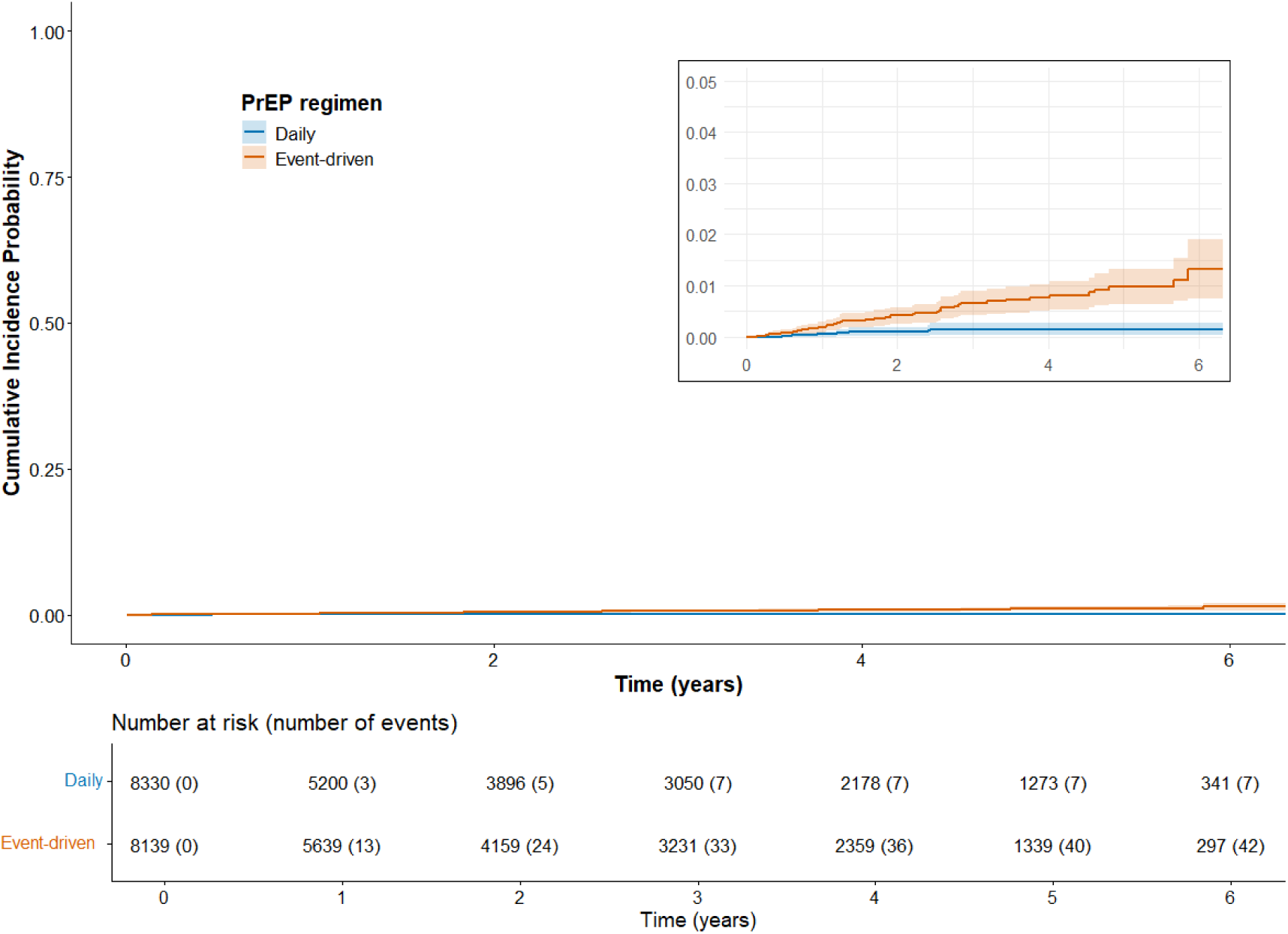
Descriptive cumulative HIV incidence curves with time-varying PrEP regimen (number of cumulative events indicated per time-point).

**Table 3.** Unadjusted and adjusted hazard ratios for the association between recent PrEP regimen used and HIV acquisition among PrEP users in the Dutch national PrEP program between August 1, 2019, and December 31, 2025.

| Characteristic | HR | 95% CI | P-value | aHR | 95% CI | P-value |
| --- | --- | --- | --- | --- | --- | --- |
| **PrEP regimen** |  |  | <0.001 |  |  | <0.001 |
| Daily | REF |  |  | REF |  |  |
| Event-driven | 5.6 | 2.5-12.5 |  | 7.0 | 3.0-16.4 |  |
| **Age(a)** | 1.0 | 1.0-1.0 | 0.50 |  |  | 0.13 |
| **Sexual preference and gender(b)** |  |  | <0.001 |  |  | 0.001 |
| MSM | REF |  |  | REF |  |  |
| Woman/MSW/TGDP/Unknown | 5.3 | 2.3-12.4 |  | 4.6 | 1.8-11.6 |  |
| **Education level** |  |  | 0.001 |  |  | 0.018 |
| Tertiary | REF |  |  | REF |  |  |
| Upper secondary/vocational | 2.2 | 1.1-4.5 |  | 2.2 | 1.0-4.4 |  |
| Lower secondary/preparatory | 2.6 | 1.0-6.5 |  | 2.1 | 0.8-5.2 |  |
| Unknown | 4.4 | 2.0-9.5 |  | 3.1 | 1.5-6.7 |  |
| **Migration background(c)** |  |  | 0.005 |  |  | 0.011 |
| No migration background | REF |  |  | REF |  |  |
| First generation migration background | 2.6 | 1.4-4.8 |  | 2.6 | 1.3-5.1 |  |
| Second generation migration background | 2.8 | 1.2-6.5 |  | 2.8 | 1.2-6.5 |  |
| **Number of partners(d)** |  |  | 0.002 |  |  | 0.004 |
| 0-4 | REF |  |  | REF |  |  |
| 5-9 | 0.6 | 0.3-1.3 |  | 0.6 | 0.3-1.2 |  |
| 10+ | 0.5 | 0.2-1.0 |  | 0.3 | 0.2-0.8 |  |
| Unknown | 5.2 | 1.6-17.7 |  | 2.7 | 0.8-10.1 |  |
| **Condomless anal sex(d)** |  |  | 0.30 |  |  | 0.30 |
| No | REF |  |  | REF |  |  |
| Yes | 3.7 | 0.5-26.7 |  | 4.5 | 0.6-35.4 |  |
| Unknown | 8.8 | 0.6-141.0 |  | 5.4 | 0.5-64.0 |  |
| **Group sex(d)** |  |  | 0.70 |  |  | 0.70 |
| No | REF |  |  | REF |  |  |
| Yes | 1.3 | 0.7-2.3 |  | 1.4 | 0.7-2.7 |  |
| Unknown | 1.3 | 0.4-4.0 |  | 0.9 | 0.3-3.0 |  |
| **Chemsex(d,e)** |  |  | 0.002 |  |  | 0.010 |
| No | REF |  |  | REF |  |  |
| Yes | 3.3 | 1.6-6.5 |  | 2.7 | 1.4-5.0 |  |
| Unknown | 2.8 | 0.4-21.1 |  | 1.1 | 0.2-5.6 |  |
| **Sex work(d)** |  |  | 0.002 |  |  | 0.063 |
| No | REF |  |  | REF |  |  |
| Yes | 3.5 | 1.4-8.8 |  | 1.8 | 0.5-5.7 |  |
| Unknown | 4.4 | 1.5-12.7 |  | 3.3 | 1.2-9.0 |  |
| **STI diagnosis in past year** |  |  | <0.001 |  |  | <0.001 |
| No/Unknown | REF |  |  | REF |  |  |
| Yes | 3.7 | 1.9-7.2 |  | 3.9 | 2.0-7.5 |  |

Note: Corresponding descriptive characteristics of the underlying Cox regression data are available in Supplementary Table S1.
(a) Age was included in the multivariable Cox model as a non-linear term using a penalized smoothing spline with four degrees of freedom.
(c) 19 missing; not included as a separate category due to zero observed events in this category.
(d) In previous six months.
(e) Defined as the use of methamphetamine, mephedrone, GHB/GBL, ketamine, cocaine, amphetamine/speed, 3-MMC, or any new psychoactive substances before or during sex.
IQR, Inter Quartile Range; MSW, Men who have Sex with Women; TGDP, Transgender and Gender Diverse Persons; MSM, Men who have Sex with Men; STI; Sexually Transmitted Infection.

## DISCUSSION

Using nationwide real-world data with over six-years of follow-up, we showed that HIV incidence among oral PrEP users was very low. However, the chance of HIV acquisition was seven-fold higher after periods of event-driven PrEP use than after periods of daily PrEP use. Given the high efficacy of both regimens and the strong association between efficacy and correct use,^1,2^ these differences are unlikely to reflect the biological efficacy of the event-driven PrEP regimen itself, but rather its effectiveness in routine practice (e.g., driven by adherence and practical application).

The higher HIV incidence during event-driven PrEP use observed in our study contrasts earlier studies, that reported similar HIV incidence in individuals using event-driven and daily PrEP, including the PREVENIR,^9,10^ the open-label phase of IPERGAY,^11^ Be-PreP-ared^12^ and AMPrEP^13^ cohort studies. Some recent studies have reported a higher HIV incidence among individuals using event-driven compared to daily PrEP, which was largely attributed to differences in adherence.^6,8^ Of note, these findings originate from West Africa and Poland and were limited by small sample sizes, short follow-up periods, and for the Poland study comparing HIV incidence was not the main study aim. Adherence is crucial for the effectiveness of both daily and event-driven PrEP, but adherence requirements differ. Whereas daily PrEP does not relate to anticipating sexual behavior and could become habitual, event-driven PrEP requires users to repeatedly and correctly initiate PrEP in accordance to anticipated sexual behavior with potential HIV exposure, and subsequently plan doses after sexual activity. Previous studies have reported challenges related to the correct initiation and adherence to the event-driven PrEP regimen, such as uncovered unplanned sex or missing post-sex doses.^14,15^ High rates of event-driven PrEP adherence have also been reported,^9,11,16^ but these high rates were observed within study settings, while feasibility of event-driven PrEP may vary in real-world settings. Although we did not have data on PrEP adherence and were therefore unable to directly assess the contribution of adherence to the observed difference in HIV incidence, lower adherence likely explains the increased HIV incidence during event-driven PrEP use. PrEP discontinuation and LTFU may also be related to barriers to PrEP use, and thus with lower adherence. We found higher rates of PrEP discontinuation and LTFU among people who most recently used event-driven PrEP. Among those seeking to re-initiate PrEP after discontinuation and LTFU, eight people were found to have acquired HIV upon re-entry, and five out of those eight HIV diagnoses were among individuals who use event-driven PrEP at their last PrEP visit. Altogether, these outcomes highlight the critical importance of person-centered guidance for those who want to use an event-driven PrEP regimen with particular attention to correct use, the possibility to switching to daily use, and HIV prevention when stopping event-driven PrEP use.

Our findings highlight the importance of counseling around PrEP choices. Event-driven PrEP has many advantages and is preferred by considerable proportions of PrEP users in various countries.^9,17–19^ It provides a more flexible and inclusive option that aligns with different user lifestyles, preferences, and changing needs.^20^ Additionally, it involves lower overall costs and is thus economically more efficient.^21^ In our study, event-driven use was chosen relative frequently, and the proportion starting event-driven PrEP increased over time. Possibly, this was related to the increased costs of PrEP tablets after August 2024. However, the event-driven regimen is likely more challenging to adhere to when compared to daily PrEP. Successful use requires anticipating sexual activity, taking tablets at multiple moments around sexual activity, and using additional prevention methods such as condoms if sexual intercourse occurs without starting PrEP. PrEP use may also be influenced by a range of individual and contextual factors, such as mood, substance use, fear of stigma, peer pressure, financial barriers, or competing life events.^22,23^ These challenges underscore the need for comprehensive and repeated counselling when initiating and during use of event-driven PrEP. Counselling should not only address dosing instructions but should also help individuals to assess whether the event-driven PrEP regimen continues to fit their lifestyle, preferences and needs and to help them anticipate and cope with challenges in correct use, including the application of other HIV prevention strategies when even-driven PrEP is not timely initiated. Furthermore, there is a need for alternative, effective, evidence-based, intermittent dosing schedules that maintain appropriate protection while decreasing the complexity of the regimen. In the United Kingdom, a 4-days-a-week regimen (TTSS, Tuesday, Thursday, Saturday, Sunday) is included in guideline.^24^ However, evidence to support effectiveness of this regime is scarce and based on post-hoc analyses.^25^

Besides choices in oral PrEP regimens, there is a need for a broader choice in HIV prevention modalities. Long-acting PrEP could provide a powerful alternative for individuals who do not want to use daily oral PrEP or those who find event-driven PrEP challenging to use. Long-acting PrEP would reduce both the burden of daily pill taking as well as the burden to plan the intake of pills around sexual activity. However, despite the high efficacy of both cabotegravir and lenacapavir as HIV prevention,^26–28^ access remains limited in many countries, including the Netherlands. High product costs and concerns about cost-effectiveness are underlying reasons for slow implementation. Expanding the variety of available HIV prevention options is increasingly important because the decline in new HIV diagnoses has plateaued in several countries including the Netherlands, even though progress has been made towards the 2030 sustainable development goals.^29^ In contexts where prevention budgets are under serious pressure, broadening the range of tailored prevention modalities ensures that interventions better align with individual preferences. Interventions can thus be tailored to the needs of diverse populations, thereby sustaining low HIV incidence and optimizing the allocation of resources.

Our findings should be interpreted considering several limitations. First, due to the observational nature of our study, we demonstrate associations rather than direct causal effects between PrEP regimen and HIV incidence. While PrEP itself is biologically protective against HIV acquisition, an individuals’ chosen regimen often serves as a proxy for underlying sexual behavioral patterns vulnerable to HIV acquisition. Although we adjusted for a range of potential confounders such as condomless anal sex and STI history, residual confounding cannot be excluded. Specifically, our surveillance data lacked information on individual adherence, psychosocial factors (e.g., mental health, planning difficulties, risk perception) and contextual factors (e.g., socio-economic status). Second, in our setting, around half of all PrEP users opt initially for event-driven use, and our outcomes thus apply to settings with a similar high interest in event-driven use. Despite these limitations, a major strength of our study is the large, real-world cohort providing over six years of data. While PrEP surveillance programs are widespread globally, few studies directly compared longitudinal HIV incidence across different PrEP regimens in routine clinical care.

In conclusion, despite an overall low HIV incidence among PrEP users in the Netherlands, we found a higher chance of acquiring HIV during periods of event-driven PrEP use than periods of daily PrEP use. While event-driven oral PrEP has been demonstrated to be as effective as daily use in study settings, most likely as a result of good adherence, individual and contextual challenges might have led to insufficient adherence resulting in lower effectiveness in a real-world setting. This underscores the need to better understand the real-world challenges relating to event driven PrEP use and improved person-centered counseling for individuals who want to start or are using event-driven PrEP.

## CONTRIBUTORS

IW, MP, EH and VJ conceptualized and designed the study. IW and VJ accessed and verified the data and were involved in the data management and analysis. EA provided support for the data analyses. IW, MP, UD, MSvdL, JH, EH and VJ were involved with interpretation of the data. IW drafted the manuscript, and EH and VJ provided substantial input and critical revisions. All authors revised and approved the final manuscript. EH and VJ had final responsibility for the decision to submit for publication.

## DATA SHARING

This study used data from the Dutch national registration of Centers for Sexual Health consultations (SOAP). Pseudonymised individual participant data can be requested for scientific use with a methodologically sound proposal submitted to the SOAP registration committee for approval. Proposal forms and additional information can be requested via. Data requestors will need to sign a data access agreement.

## ACKNOWLEDGEMENTS

All data on PrEP use and HIV diagnoses used in these analyses are obtained from the national STI surveillance database SOAP at the RIVM, collected through all 23 Centers for Sexual Health in the Netherlands. During the preparation of this work, the authors used generative AI (Gemini and Mistral) to improve the language, readability, and flow of the text. The authors reviewed and edited the content and take full responsibility for the final manuscript.

## DECLARATION OF GENERATIVE AI AND AI-ASSISTED TECHNOLOGIES IN THE MANUSCRIPT PREPARATION PROCESS

During the preparation of this work, the authors used Gemini, Mistral and ChatGPT for coding assistance, brainstorming data analysis approaches, and refining the text for clarity and readability. The authors reviewed and edited the output as needed and take full responsibility for the content of the published article.

