## Supplemental Files for "Increased HIV incidence during Event-Driven PrEP compared to Daily PrEP in the Netherlands"

**SUPPLEMENTARY MATERIALS - CONTENTS**

**Supplementary Table S1.** Descriptive characteristics at PrEP visit level, stratified by HIV acquisition during follow-up (Dutch national PrEP program, Aug2019–Dec2025).

**Supplementary Figure S2.** Descriptive cumulative HIV incidence curves with time-varying PrEP regimen reported in the past year (number of cumulative events indicated per time-point).

**Supplementary Table S3.** Unadjusted and adjusted hazard ratios for the association between PrEP regimen in the past year and HIV acquisition among PrEP users in the Dutch National PrEP Program between August 1, 2019, and December 31, 2025.

**Supplementary Table S4.** Unadjusted hazard ratios of interaction terms between covariates and PrEP regimen on the association between PrEP regimen and HIV acquisition among PrEP users in the Dutch National PrEP Program between 2019 and 2025.

**Supplementary Material S5.** Missing Data Analysis

**Supplementary Figure S5.1**. Descriptive cumulative HIV incidence curves with time-varying PrEP regimen, using complete case analysis (number of cumulative events indicated per time-point).

**Supplementary Table S5.2.** Unadjusted and adjusted hazard ratios for the association between PrEP regimen and HIV acquisition among PrEP users in the Dutch National PrEP Program between August 1, 2019, and December 31, 2025, using complete case analysis.

**Supplementary Figure S6.** Descriptive cumulative HIV incidence curves with time-varying PrEP regimen: sensitivity analysis assuming all individuals initiated PrEP using a daily regimen (number of cumulative events indicated per time-point).

**Supplementary Table S7.** Unadjusted and adjusted hazard ratios for the association between PrEP regimen and HIV acquisition among PrEP users in the Dutch national PrEP program between August 1, 2019, and December 31, 2025: sensitivity analysis assuming all individuals initiated PrEP using a daily regimen.

**Supplementary Table S1. Descriptive characteristics at visit level (including both STI and PrEP visits), stratified by HIV acquisition during follow-up (Dutch national PrEP program, Aug2019–Dec2025).**

| **Characteristic** | **Overall  N = 144,927** | **No HIV diagnosis  N = 144,878** | **HIV diagnosis  N = 49** |
| --- | --- | --- | --- |
| **PrEP regimen reported in past three months** |  |  |  |
| Daily | 75,471 (52.1%) | 75,464 (52.1%) | 7 (14.3%) |
| Event-driven | 69,456 (47.9%) | 69,414 (47.9%) | 42 (85.7%) |
| **Type of visit** |  |  |  |
| STI visit | 21,422 (14.8%) | 21,405 (14.8%) | 17 (34.7%) |
| PrEP intiation visit | 555 (0.4%) | 555 (0.4%) | 0 (0.0%) |
| PrEP follow-up visit | 122,950 (84.8%) | 122,918 (84.8%) | 32 (65.3%) |
| **Age (years), median[IQR]** | 37 (30 - 48) | 37 (30 - 48) | 33 (29 - 48) |
| **Sexual preference and gender(a)** |  |  |  |
| MSM | 141,192 (97.4%) | 141,149 (97.4%) | 43 (87.8%) |
| Woman/MSW/TGDP/Unknown | 3,735 (2.6%) | 3,729 (2.6%) | 6 (12.2%) |
| **Education level** |  |  |  |
| Tertiary | 92,506 (63.8%) | 92,487 (63.8%) | 19 (38.8%) |
| Upper secondary/vocational | 30,024 (20.7%) | 30,010 (20.7%) | 14 (28.6%) |
| Lower secondary/preparatory | 11,234 (7.8%) | 11,228 (7.7%) | 6 (12.2%) |
| Unknown | 11,163 (7.7%) | 11,153 (7.7%) | 10 (20.4%) |
| **Migration background(b)** |  |  |  |
| No migration background | 82,847 (57.2%) | 82,830 (57.2%) | 17 (34.7%) |
| First generation migration background | 47,682 (32.9%) | 47,658 (32.9%) | 24 (49.0%) |
| Second generation migration background | 14,398 (9.9%) | 14,390 (9.9%) | 8 (16.3%) |
| **Number of partners(c)** |  |  |  |
| 0-4 | 45,967 (31.7%) | 45,945 (31.7%) | 22 (44.9%) |
| 5-9 | 37,954 (26.2%) | 37,943 (26.2%) | 11 (22.4%) |
| 10+ | 59,760 (41.2%) | 59,747 (41.2%) | 13 (26.5%) |
| Unknown | 1,246 (0.9%) | 1,243 (0.9%) | 3 (6.1%) |
| **Condomless anal sex(c)** |  |  |  |
| No | 9,787 (6.8%) | 9,786 (6.8%) | 1 (2.0%) |
| Yes | 133,869 (92.4%) | 133,822 (92.4%) | 47 (95.9%) |
| Unknown | 1,271 (0.9%) | 1,270 (0.9%) | 1 (2.0%) |
| **Group sex(c)** |  |  |  |
| No | 71,813 (49.6%) | 71,791 (49.6%) | 22 (44.9%) |
| Yes | 62,418 (43.1%) | 62,395 (43.1%) | 23 (46.9%) |
| Unknown | 10,696 (7.4%) | 10,692 (7.4%) | 4 (8.2%) |
| **Chemsex(c,d)** |  |  |  |
| No | 123,547 (85.2%) | 123,515 (85.3%) | 32 (65.3%) |
| Yes | 20,104 (13.9%) | 20,088 (13.9%) | 16 (32.7%) |
| Unknown | 1,276 (0.9%) | 1,275 (0.9%) | 1 (2.0%) |
| **Sex work(c)** |  |  |  |
| No | 136,215 (94.0%) | 136,175 (94.0%) | 40 (81.6%) |
| Yes | 5,315 (3.7%) | 5,310 (3.7%) | 5 (10.2%) |
| Unknown | 3,397 (2.3%) | 3,393 (2.3%) | 4 (8.2%) |
| **STI diagnosis in past year** |  |  |  |
| No/Unknown | 68,923 (47.6%) | 68,912 (47.6%) | 11 (22.4%) |
| Yes | 76,004 (52.4%) | 75,966 (52.4%) | 38 (77.6%) |
| Note: Data are presented at the consultation/visit level (N represents total person-time observations, not unique individuals) to reflect the data underlying the Cox regression model.  (a) Gender/sexual preference was analyzed as time-varying variable to account for changes during the follow-up period, which results in a different distribution in the baseline characteristics presented in Table 1.  (b) 19 missing; not included as a separate category due to zero observed events in this category. (c) In previous six months.  (d) Defined as the use of methamphetamine, mephedrone, GHB/GBL, ketamine, cocaine, amphetamine/speed, 3-MMC, or any new psychoactive substances before or during sex.  IQR, Inter Quartile Range; MSW, Men who have Sex with Women; Transgender and Gender Diverse Persons; MSM, Men who have Sex with Men; STI; Sexually Transmitted Infection. | | | |

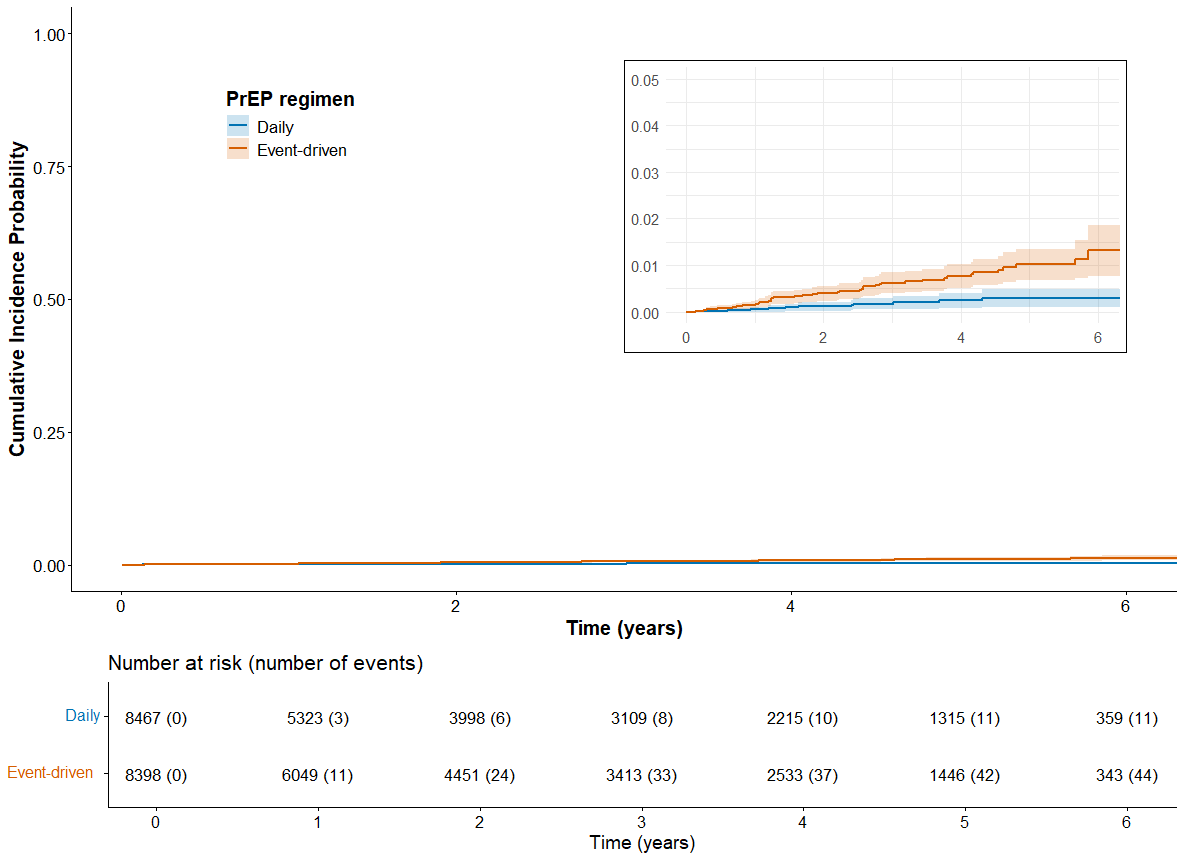

**Supplementary Figure S2**. **Descriptive cumulative HIV incidence curves with time-varying PrEP regimen reported in the past year (number of cumulative events indicated per time-point).**

**Supplementary Table S3. Unadjusted and adjusted hazard ratios for the association between PrEP regimen in the past year and HIV acquisition among PrEP users in the Dutch National PrEP Program between August 1, 2019, and December 31, 2025.**

| **Characteristic** | **HR** | **95% CI** | **P-value** | **aHR** | **95% CI** | **P-value** |
| --- | --- | --- | --- | --- | --- | --- |
| **PrEP regimen** |  |  | <0.001 |  |  | <0.001 |
| Daily | REF | |  | REF | |  |
| Event-driven | 3.6 | 1.9-7.0 |  | 4.3 | 2.1-8.7 |  |
| **Age(a)** | 1.0 | 1.0-1.0 | 0.20 |  |  | 0.20 |
| **Sexual preference and gender** |  |  | <0.001 |  |  | <0.001 |
| MSM | REF | |  | REF | |  |
| Woman/MSW/TGDP/Unknown | 5.4 | 2.4-11.9 |  | 4.8 | 2.0-11.6 |  |
| **Education level** |  |  | 0.001 |  |  | 0.013 |
| Tertiary | REF | |  | REF | |  |
| Upper secondary/vocational | 2.6 | 1.4-4.8 |  | 2.5 | 1.3-4.9 |  |
| Lower secondary/preparatory | 2.3 | 0.9-5.8 |  | 1.8 | 0.8-4.4 |  |
| Unknown | 4.0 | 1.9-8.4 |  | 2.8 | 1.3-5.9 |  |
| **Migration background(b)** |  |  | <0.001 |  |  | 0.002 |
| No migration background | REF | |  | REF | |  |
| First generation migration background | 2.8 | 1.5-5.0 |  | 2.7 | 1.4-5.2 |  |
| Second generation migration background | 3.3 | 1.5-7.2 |  | 3.1 | 1.4-6.8 |  |
| **Number of partners(c)** |  |  | <0.001 |  |  | 0.001 |
| 0-4 | REF | |  | REF | |  |
| 5-9 | 0.5 | 0.3-1.1 |  | 0.5 | 0.2-1.0 |  |
| 10+ | 0.4 | 0.2-0.9 |  | 0.3 | 0.1-0.7 |  |
| Unknown | 4.5 | 1.3-14.8 |  | 2.6 | 0.7-9.3 |  |
| **Condomless anal sex(c)** |  |  | 0.50 |  |  | 0.50 |
| No | REF | |  | REF | |  |
| Yes | 2.1 | 0.5-8.6 |  | 2.5 | 0.6-11.1 |  |
| Unknown | 3.8 | 0.3-41.9 |  | 2.4 | 0.3-17.9 |  |
| **Group sex(c)** |  |  | >0.9 |  |  | 0.80 |
| No | REF | |  | REF | |  |
| Yes | 1.1 | 0.6-1.9 |  | 1.2 | 0.6-2.4 |  |
| Unknown | 1.1 | 0.4-3.4 |  | 0.8 | 0.3-2.5 |  |
| **Chemsex(c,d)** |  |  | 0.004 |  |  | 0.007 |
| No | REF | |  | REF | |  |
| Yes | 3.0 | 1.6-5.7 |  | 2.6 | 1.4-4.9 |  |
| Unknown | 2.3 | 0.3-17.1 |  | 1.0 | 0.2-4.6 |  |
| **Sex work(c)** |  |  | 0.005 |  |  | 0.12 |
| No | REF | |  | REF | |  |
| Yes | 3.0 | 1.2-7.6 |  | 1.3 | 0.4-4.2 |  |
| Unknown | 3.8 | 1.3-10.9 |  | 2.8 | 1.0-7.8 |  |
| **STI diagnosis in past year** |  |  | <0.001 |  |  | <0.001 |
| No/Unknown | REF | |  | REF | |  |
| Yes | 4.4 | 2.2-8.5 |  | 4.7 | 2.4-9.1 |  |
| (a) Age was included in the multivariable Cox model as a non-linear term using a penalized smoothing spline with four degrees of freedom. (b) 19 missing; not included as a separate category due to zero observed events in this category. (c) In previous six months.  (d) Defined as the use of methamphetamine, mephedrone, GHB/GBL, ketamine, cocaine, amphetamine/speed, 3-MMC, or any new psychoactive substances before or during sex.  IQR, Inter Quartile Range; MSW, Men who have Sex with Women; TGDP, Transgender and Gender Diverse Persons; MSM, Men who have Sex with Men; STI; Sexually Transmitted Infection. | | | | | | |

**Supplementary Table S4. Unadjusted hazard ratios of interaction terms between covariates and PrEP regimen on the association between PrEP regimen and HIV acquisition among PrEP users in the Dutch National PrEP Program between 2019 and 2025.**

| **Covariate** | **Interaction Term** | **Hazard Ratio (HR)** | **95% CI** | **P-value** |
| --- | --- | --- | --- | --- |
| Group sex | PrEP regimen × Yes | 0.3 | [0.0; 2.6] | 0.255 |
| Group sex | PrEP regimen × Unknown | 0.1 | [0.0; 1.4] | 0.081 |
| Number of partners | PrEP regimen × 5-9 | 1.2 | [0.1; 15.0] | 0.879 |
| Number of partners | PrEP regimen × 10+ | 0.7 | [0.1; 5.0] | 0.733 |
| Number of partners | PrEP regimen × Unknown | 0.3 | [0.0; 5.7] | 0.454 |
| Sexual preference and gender | PrEP regimen × Woman/MSW/TGDP/Unknown | 0.6 | [0.1; 4.5] | 0.660 |
| Sex work | PrEP regimen × Yes | 1.8 | [0.2; 18.9] | 0.634 |
| Sex work | PrEP regimen × Unknown | NE | NE | NE |
| Education level | PrEP regimen × Upper secondary/vocational | 0.7 | [0.1; 6.0] | 0.779 |
| Education level | PrEP regimen × Lower secondary/preparatoy | 0.7 | [0.1; 9.9] | 0.817 |
| Education level | PrEP regimen × Unknown | 0.7 | [0.1; 5.6] | 0.709 |
| Migration background | PrEP regimen × First generation migration background | 1.9 | [0.2; 14.7] | 0.556 |
| Migration background | PrEP regimen × Second generation migration background | 0.2 | [0.0; 1.6] | 0.129 |
| Age | PrEP regimen × Age | 1.0 | [0.9; 1.1] | 0.947 |
| Condomless anal sex | PrEP regimen × Yes | NE | NE | NE |
| Condomless anal sex | PrEP regimen × Unknown | NE | NE | NE |
| Chemsex | PrEP regimen × Yes | 1.0 | [0.2; 6.1] | 0.960 |
| Chemsex | PrEP regimen × Unknown | NE | NE | NE |
| STI diagnosis in past year | PrEP regimen × Yes | 0.8 | [0.1; 7.2] | 0.822 |

STI; Sexually Transmitted Infection. NE: Not estimable due to low event rate in this subgroup.

**Supplementary Material S5.** Missing Data Analysis

Sensitivity Analyses for Handling Missing Data

To assess the impact of missing data handling on our primary outcomes, two sensitivity analyses were performed:

1) Direction of Imputation (LOCF vs. LOCB): We compared Last Observation Carried Forward (LOCF) with Last Observation Carried Backward (LOCB). Both methods yielded virtually identical results; ten missed consultations were imputed as daily PrEP (prepreg == 0) and five as event-driven PrEP (prepreg == 1), with only one consultation remaining unfulfilled in the LOCB approach. Because imputation direction did not affect the estimates, full LOCB results are not shown.
2) Complete Case Analysis: Additionally, a complete case analysis was conducted as an alternative to imputation. Descriptive cumulative HIV incidence curves based on complete cases are presented in Supplementary Figure S5.1.

Complete case analysis

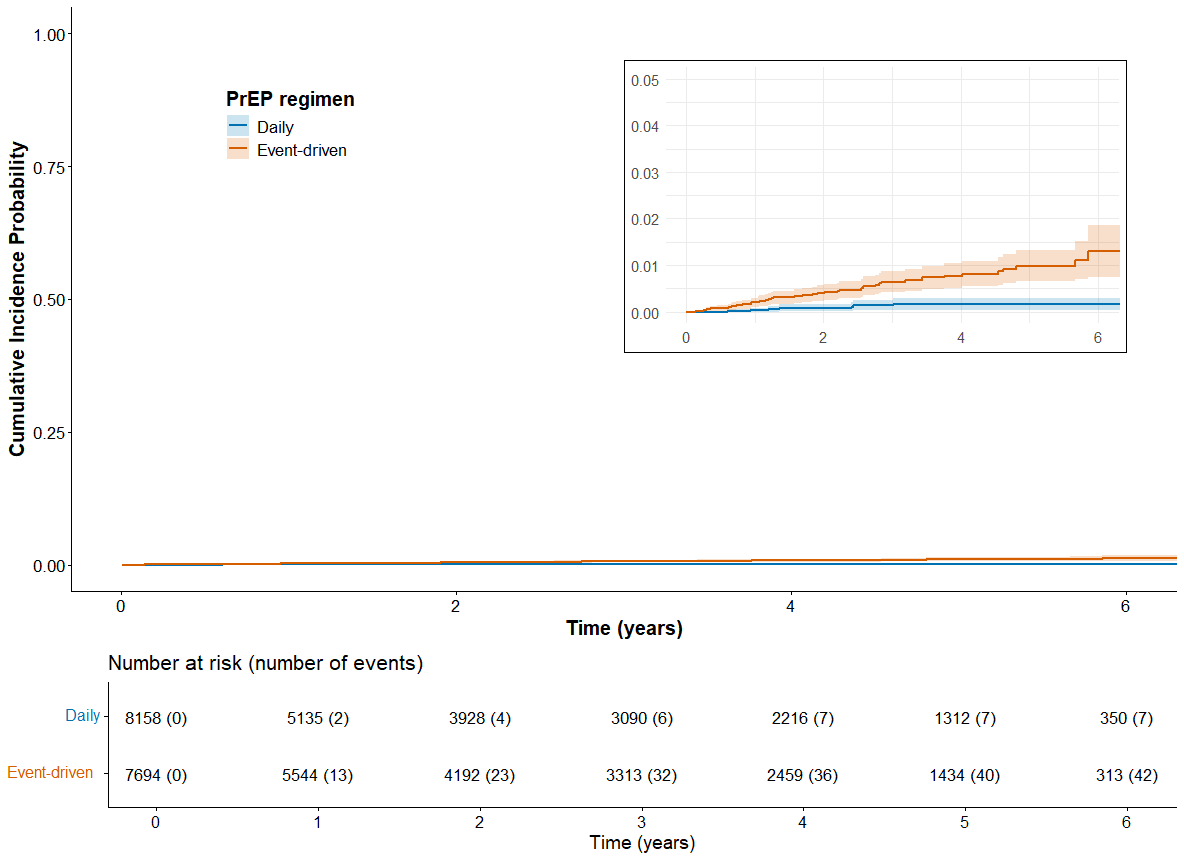

**Supplementary Figure S5.1**. **Descriptive cumulative HIV incidence curves with time-varying PrEP regimen, using complete case analysis (number of cumulative events indicated per time-point).**

Note: this analysis excludes the 15 consultations with missing PrEP regimen data (which were imputed as ten daily and five event-driven PrEP in the primary analysis).

**Supplementary Table S5.2. Unadjusted and adjusted hazard ratios for the association between PrEP regimen and HIV acquisition among PrEP users in the Dutch National PrEP Program between August 1, 2019, and December 31, 2025, using complete case analysis.**

| **Characteristic** | **HR** | **95% CI** | **P-value** | **aHR** | **95% CI** | **P-value** |
| --- | --- | --- | --- | --- | --- | --- |
| **PrEP regimen** |  |  | <0.001 |  |  | <0.001 |
| Daily | REF | |  | REF | |  |
| Event-driven | 5.6 | 2.5-12.5 |  | 7.0 | 3.0-16.4 |  |
| **Age(a)** | 1.0 | 1.0-1.0 | 0.40 |  |  | 0.14 |
| **Sexual preference and gender** |  |  | <0.001 |  |  | 0.001 |
| MSM | REF | |  | REF | |  |
| Woman/MSW/TGDP/Unknown | 5.3 | 2.3-12.5 |  | 4.5 | 1.8-11.3 |  |
| **Education level** |  |  | 0.001 |  |  | 0.017 |
| Tertiary | REF | |  | REF | |  |
| Upper secondary/vocational | 2.2 | 1.1-4.4 |  | 2.2 | 1.0-4.5 |  |
| Lower secondary/preparatory | 2.6 | 1.0-6.5 |  | 2.1 | 0.8-5.2 |  |
| Unknown | 4.4 | 2.1-9.5 |  | 3.1 | 1.5-6.8 |  |
| **Migration background(b)** |  |  | 0.005 |  |  | 0.011 |
| No migration background | REF | |  | REF | |  |
| First generation migration background | 2.6 | 1.4-4.8 |  | 2.8 | 1.2-6.5 |  |
| Second generation migration background | 2.8 | 1.2-6.5 |  | 2.5 | 1.3-5.1 |  |
| **Number of partners(c)** |  |  | 0.002 |  |  | 0.004 |
| 0-4 | REF | |  | REF | |  |
| 5-9 | 0.6 | 0.3-1.3 |  | 0.6 | 0.3-1.2 |  |
| 10+ | 0.5 | 0.2-1.0 |  | 0.3 | 0.2-0.8 |  |
| Unknown | 5.2 | 1.5-17.5 |  | 2.8 | 0.8-10.1 |  |
| **Condomless anal sex(c)** |  |  | 0.30 |  |  | 0.40 |
| No | REF | |  | REF | |  |
| Yes | 3.7 | 0.5-27.0 |  | 4.4 | 0.6-35.4 |  |
| Unknown | 8.8 | 0.6-140.0 |  | 5.2 | 0.4-64.6 |  |
| **Group sex(c)** |  |  | 0.70 |  |  | 0.70 |
| No | REF | |  | REF | |  |
| Yes | 1.3 | 0.7-2.3 |  | 1.3 | 0.7-2.7 |  |
| Unknown | 1.4 | 0.4-4.1 |  | 1.0 | 0.3-3.1 |  |
| **Chemsex(c,d)** |  |  | 0.003 |  |  | 0.010 |
| No | REF | |  | REF | |  |
| Yes | 3.2 | 1.6-6.4 |  | 2.7 | 1.4-5.1 |  |
| Unknown | 2.9 | 0.4-21.3 |  | 1.1 | 0.2-5.7 |  |
| **Sex work(c)** |  |  | 0.001 |  |  | 0.055 |
| No | REF | |  | REF | |  |
| Yes | 3.5 | 1.4-8.9 |  | 1.8 | 0.6-5.7 |  |
| Unknown | 4.4 | 1.5-13.0 |  | 3.3 | 1.2-9.2 |  |
| **STI diagnosis in past year** |  |  | <0.001 |  |  | <0.001 |
| No/Unknown | REF | |  | REF | |  |
| Yes | 3.7 | 1.9-7.2 |  | 3.9 | 2.0-7.5 |  |
| Note: this analysis excludes the 15 consultations with missing PrEP regimen data (which were imputed as ten daily and five event-driven PrEP in the primary analysis). (a) Age was included in the multivariable Cox model as a non-linear term using a penalized smoothing spline with four degrees of freedom. (b) 19 missing; not included as a separate category due to zero observed events in this category. (c) In previous six months.  (d) Defined as the use of methamphetamine, mephedrone, GHB/GBL, ketamine, cocaine, amphetamine/speed, 3-MMC, or any new psychoactive substances before or during sex.  IQR, Inter Quartile Range; MSW, Men who have Sex with Women; TGDP, Transgender and Gender Diverse Persons; MSM, Men who have Sex with Men; STI; Sexually Transmitted Infection. | | | | | | |

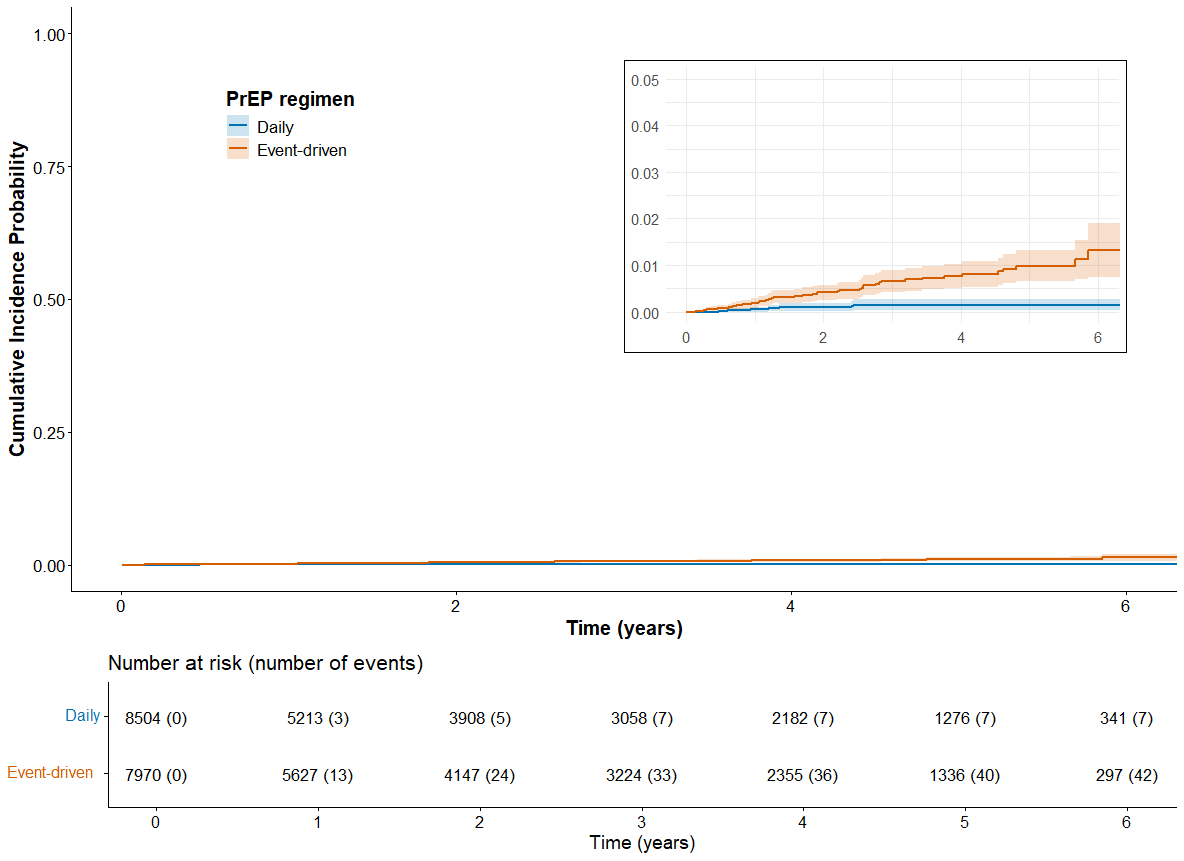

**Supplementary Figure S6. Descriptive cumulative HIV incidence curves with time-varying PrEP regimen: sensitivity analysis assuming all individuals initiated PrEP using a daily regimen (number of cumulative events indicated per time-point).**

**Supplementary Table S7. Unadjusted and adjusted hazard ratios for the association between PrEP regimen and HIV acquisition among PrEP users in the Dutch national PrEP program between August 1, 2019, and December 31, 2025: sensitivity analysis assuming all individuals initiated PrEP using a daily regimen.**

| **Characteristic** | **HR** | **95% CI** | **P-value** | **aHR** | **95% CI** | **P-value** |
| --- | --- | --- | --- | --- | --- | --- |
| **PrEP regimen** |  |  | <0.001 |  |  | <0.001 |
| Daily | REF | |  | REF | |  |
| Event-driven | 5.7 | 2.5-12.6 |  | 7.1 | 3.0-16.6 |  |
| **Age(a)** | 1.0 | 1.0-1.0 | 0.50 |  |  | 0.13 |
| **Sexual preference and gender** |  |  | <0.001 |  |  | 0.001 |
| MSM | REF | |  | REF | |  |
| Woman/MSW/TGDP/Unknown | 5.3 | 2.3-12.4 |  | 4.6 | 1.8-11.6 |  |
| **Education level** |  |  | 0.001 |  |  | 0.018 |
| Tertiary | REF | |  | REF | |  |
| Upper secondary/vocational | 2.2 | 1.1-4.5 |  | 2.2 | 1.0-4.4 |  |
| Lower secondary/preparatory | 2.6 | 1.0-6.5 |  | 2.1 | 0.8-5.2 |  |
| Unknown | 4.4 | 2.0-9.5 |  | 3.1 | 1.5-6.8 |  |
| **Migration background(b)** |  |  | 0.005 |  |  | 0.011 |
| No migration background | REF | |  | REF | |  |
| First generation migration background | 2.6 | 1.4-4.8 |  | 2.8 | 1.2-6.5 |  |
| Second generation migration background | 2.8 | 1.2-6.5 |  | 2.5 | 1.3-5.1 |  |
| **Number of partners(c)** |  |  | 0.002 |  |  | 0.004 |
| 0-4 | REF | |  | REF | |  |
| 5-9 | 0.6 | 0.3-1.3 |  | 0.6 | 0.3-1.2 |  |
| 10+ | 0.5 | 0.2-1.0 |  | 0.3 | 0.2-0.8 |  |
| Unknown | 5.2 | 1.6-17.7 |  | 2.8 | 0.8-10.2 |  |
| **Condomless anal sex(c)** |  |  | 0.30 |  |  | 0.30 |
| No | REF | |  | REF | |  |
| Yes | 3.7 | 0.5-26.7 |  | 4.5 | 0.6-35.6 |  |
| Unknown | 8.8 | 0.6-141.0 |  | 5.4 | 0.5-64.6 |  |
| **Group sex(c)** |  |  | 0.70 |  |  | 0.70 |
| No | REF | |  | REF | |  |
| Yes | 1.3 | 0.7-2.3 |  | 1.4 | 0.7-2.7 |  |
| Unknown | 1.3 | 0.4-4.0 |  | 0.9 | 0.3-3.0 |  |
| **Chemsex(c,d)** |  |  | 0.002 |  |  | 0.010 |
| No | REF | |  | REF | |  |
| Yes | 3.3 | 1.6-6.5 |  | 2.7 | 1.4-5.0 |  |
| Unknown | 2.8 | 0.4-21.1 |  | 1.1 | 0.2-5.6 |  |
| **Sex work(c)** |  |  | 0.002 |  |  | 0.063 |
| No | REF | |  | REF | |  |
| Yes | 3.5 | 1.4-8.8 |  | 1.8 | 0.6-5.8 |  |
| Unknown | 4.4 | 1.5-12.7 |  | 3.3 | 1.2-9.0 |  |
| **STI diagnosis in past year** |  |  | <0.001 |  |  | <0.001 |
| No/Unknown | REF | |  | REF | |  |
| Yes | 3.7 | 1.9-7.2 |  | 3.8 | 2.0-7.5 |  |
| (a) Age was included in the multivariable Cox model as a non-linear term using a penalized smoothing spline with four degrees of freedom. (b) 19 missing; not included as a separate category due to zero observed events in this category. (c) In previous six months.  (d) Defined as the use of methamphetamine, mephedrone, GHB/GBL, ketamine, cocaine, amphetamine/speed, 3-MMC, or any new psychoactive substances before or during sex.  IQR, Inter Quartile Range; MSW, Men who have Sex with Women; TGDP, Transgender and Gender Diverse Persons; MSM, Men who have Sex with Men; STI; Sexually Transmitted Infection. | | | | | | |
